# Candidate molecular predictors of outcome after aneurysmal subarachnoid haemorrhage: a systematic review of haemoglobin metabolism, inflammation and oxidative injury pathways

**DOI:** 10.1101/19004853

**Authors:** Ben Gaastra, Ian Galea

## Abstract

Aneurysmal subarachnoid haemorrhage (aSAH) is a devastating form of stroke associated with significant morbidity and mortality. Very little is known about the predictors of poor outcome and the pathophysiological mechanisms underlying neurological injury following aSAH. Three molecular pathways have been shown to be important: haemoglobin metabolism, inflammation and oxidative injury. The aim of this review is to use a systematic approach to identify a panel of key genes within these three pathways in order to focus future studies investigating predictors of poor outcome and the mechanisms of neurological injury following aSAH. Manual searching and bioinformatic mining tools were used. Studies of experimental or human SAH were included, and outcome was broadly defined to include all encountered readouts such as mortality, neurological scores, and neuropathological markers of tissue damage. If two or more molecules belonged to the same biochemical pathway, this pathway was examined in detail to identify all its components, which were then searched individually for any evidence of association with outcome using the same broad definition as before. This resulted in the identification of 58 candidate genes within the three pathways of interest (haemoglobin metabolism, inflammation and oxidative injury) potentially linked to outcome after aSAH.

## Introduction

Aneurysmal subarachnoid haemorrhage (aSAH) results from the release of blood into the subarachnoid space as a consequence of a cerebral artery aneurysm rupture. aSAH is associated with significant morbidity and mortality. Twenty-eight day mortality rates of up to 42% have been reported^1^ with the largest proportion of patients dying within two days of ictus^2^. Long term morbidity affecting survivors is significant with around half of survivors having cognitive impairments and only a third returning to the same work as prior to haemorrhage^3^. The socioeconomic burden of aSAH in the UK has been estimated as £510 million annually^4^.

Neurological injury following aSAH can be broadly considered in two categories: firstly, an early brain injury (EBI) occurring within the first 72 hours of ictus, followed by a delayed brain injury occurring days to weeks after injury. EBI is thought to occur as a consequence of a spike in intracranial pressure and an associated fall in cerebral blood flow. This initiates toxic cascades causing global cerebral ischemia, blood-brain-barrier (BBB) disruption, cerebral oedema and ultimately cell death^5^. The toxic cascades initiated by EBI and the presence of blood and its breakdown products in the subarachnoid space are thought to cause a delayed brain injury characterised by spreading cortical depression, loss of autoregulation, microthrombi, inflammation, oxidative stress and vasospasm^6-8^.

Blood within the subarachnoid space is broken down and cleared in a multi-step pathway involving a number of scavenging molecules^9^. This clearance system within the central nervous system (CNS) is not as well developed compared to the rest of the body meaning that blood break down products within the CNS persist for a relatively long time causing inflammation and oxidative stress. Blood breakdown product scavenging molecules have been shown to play an important role in mitigating toxicity and limiting the neurological injury caused by blood breakdown products. Several authoritative reviews^9-15^ have concluded that haemoglobin metabolism, inflammatory and oxidative mechanisms play a significant role in neurological injury following SAH (Figure 1).

**Figure 1.**
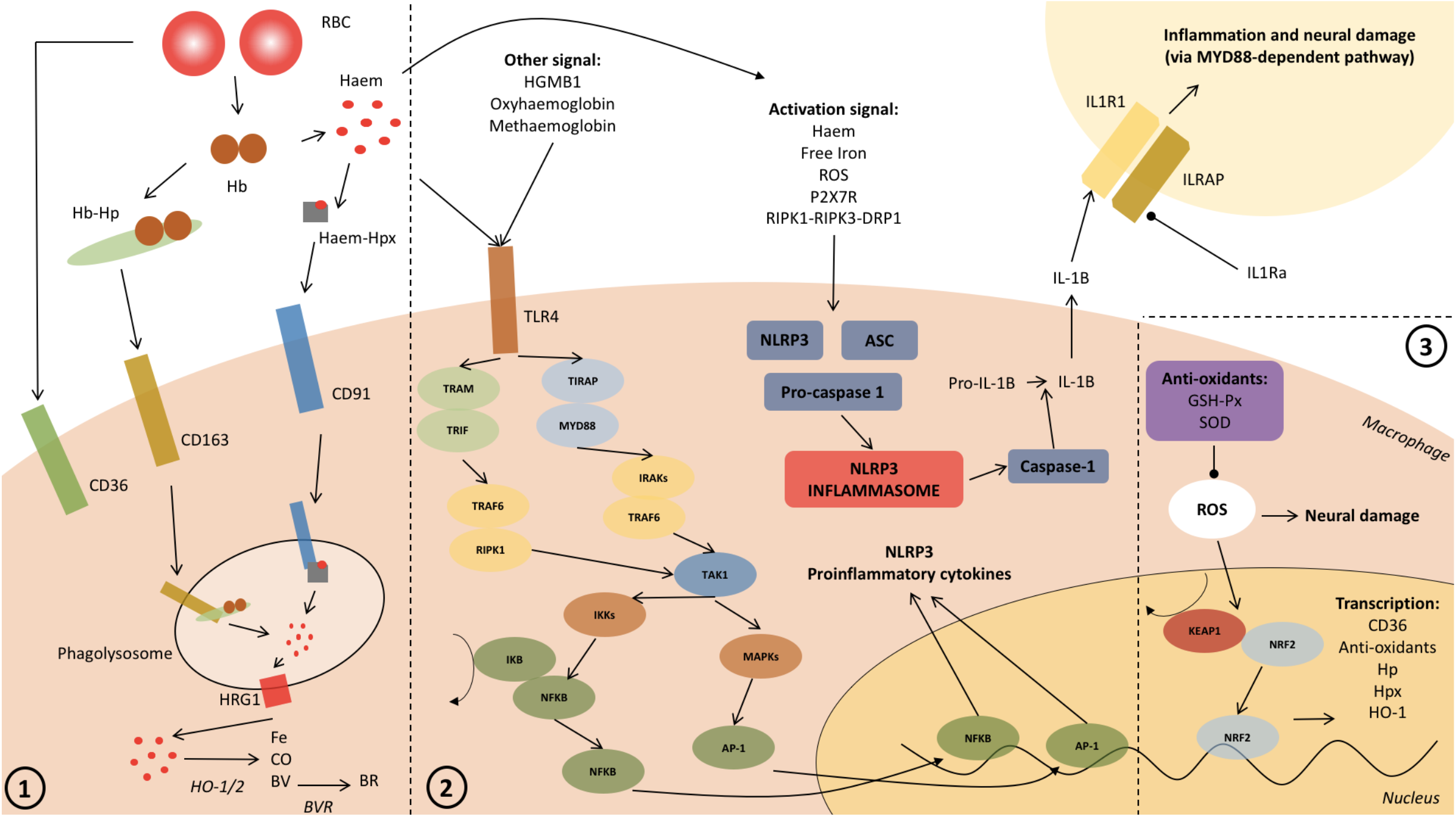
Schematic summarising blood breakdown (1), inflammatory (2) and oxidative (3) pathways involved in SAH.

The aim of this review is to identify a panel of molecules within the three themes of haemoglobin metabolism, inflammation and oxidative stress, which are of potential prognostic value after human aSAH. Candidate molecules were identified by an exhaustive search of the literature, using a combination of manual and text-mining methodology.

The candidate molecules identified here will be of use to any investigator who plans to perform a targeted study of the three pathways and outcome after SAH, using genetic, proteomic or metabolomic methods. As an example, in our case, we plan to perform a gene exome sequencing study, during which the exons and upstream regions of the candidates will be sequenced; the association of genetic variation in these regions with clinical outcome after aSAH will be investigated. Further study of these pathways will: (1) provide further evidence of the importance of haemoglobin breakdown product metabolism, inflammation and oxidative stress after human SAH; (2) improve prognostic tools; (3) give insight into the mechanisms of neurological injury following aSAH; (4) identify therapeutic targets for aSAH.

## Methods

### Rationale

A combination of manual and semi-automated methodology was used, rather than fully automated methods. In order to illustrate the need for this combined approach, we give an example. In mouse and rat models of SAH, two studies^16,17^ compared RNA expression in cortical tissue from SAH versus control animals, and found that inflammation is a key theme. Blood breakdown products and oxidative stress are intricately linked to inflammation^18-23^, yet these molecules are less likely to feature in automated pathway or network analyses of unbiased proteomic and RNA expression datasets since they are not well described in terms of KEGG pathways and GO ontology terms, and their networks are composed of a small number of molecules. For instance, when the 324 genes identified to be related to SAH by GLAD4U^24^ (a PubMed gene retrieval and prioritization tool) are run through Over-Representation Analysis (ORA), Gene Set Enrichment Analysis (GSEA), and Network Topology-based Analysis (NTA) bioinformatic pipelines, oxidative stress and blood breakdown product metabolism are not identified. Hence enrichment of potential candidates within these pathways, based on most likely biologic targets reported in the literature, is needed to study components of these pathways which have been shown to be important in determining outcome after experimental or human SAH.

### Overview

A literature search was performed for molecules shown to be important in influencing outcome after experimental or human aSAH within the three themes of haemoglobin metabolism, inflammation and oxidative stress. Outcome was broadly defined and included all encountered readouts such as mortality, neurological scores, and neuropathological markers of tissue damage.

The search strategy consisted of a manual Pubmed search specifically addressing the three themes of haemoglobin metabolism, inflammation and oxidative stress, supplemented by the full-text literature mining gene retrieval and functional analysis tool SciMiner^25^ (http://hurlab.med.und.edu/SciMiner/).

Publications retrieved were manually screened to ensure that the molecule was positively associated with outcome after SAH. For the reasons explained under “ Rationale”, a combination of literature and pathway analysis was used in a sequential multi-stage process, which is described next.

### Description of stages

The stages of this search are depicted graphically in Figure 2. First, a primary literature search was performed in order to identify molecules (referred to as ‘Group A’ molecules) relevant to SAH from the three key themes: blood breakdown product metabolism, inflammation and oxidation. PubMed was searched for articles published in English prior to January 2019 using the following criteria: “ oxidation”, “ anti-oxidant”, “ oxidative”, “ redox”, “ inflammation”, “ inflammatory”, “ h(a)emoglobin”, “ haem/heme” and “ iron” in single combination with “ subarachnoid h(a)emorrhage”. The reference lists of published articles were manually searched for further articles. The results were manually curated to select those molecules related to SAH outcome.

**Figure 2.**
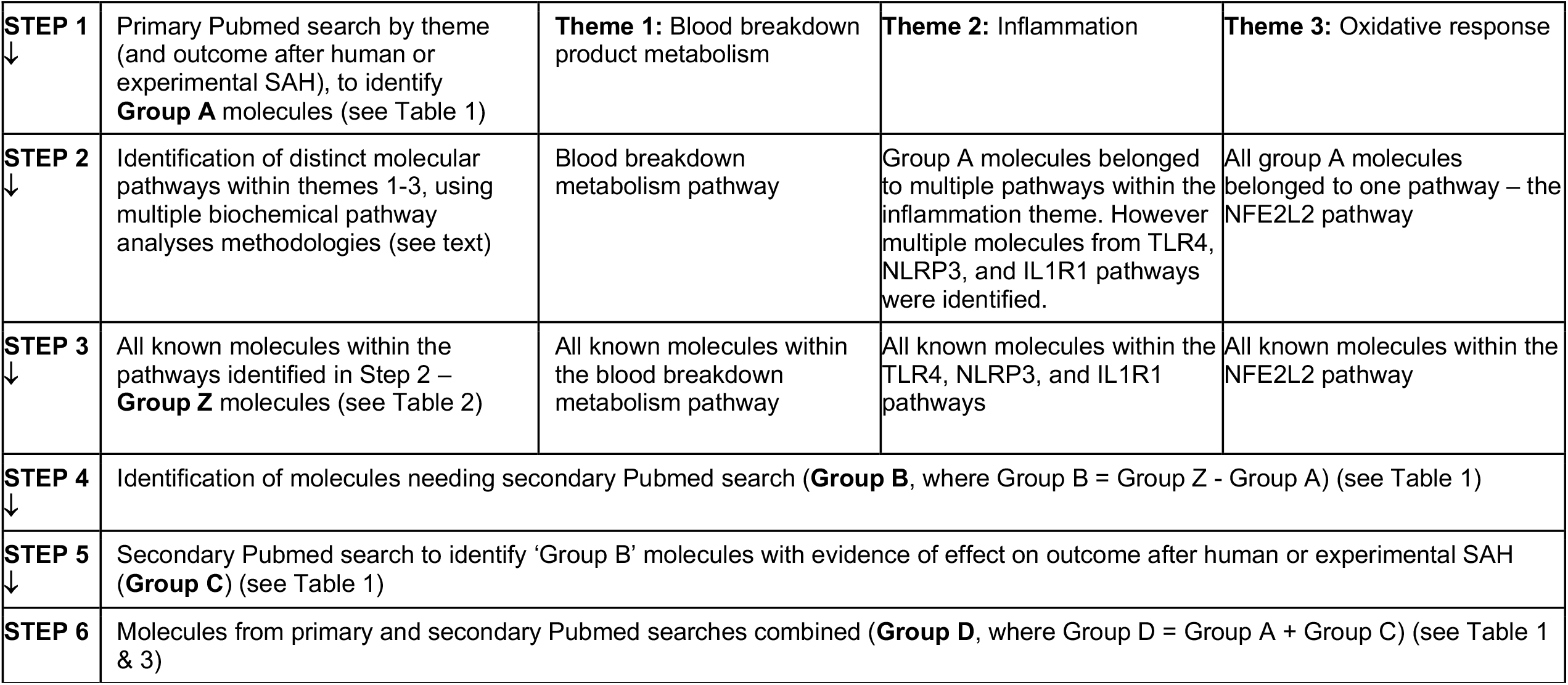
Methods flowchart

If two or more molecules identified during the primary search belonged to the same biochemical pathway, this pathway was examined using different pathway analysis tools to identify all its components: the Reactome pathway database^26^, KEGG pathway repository^27^, textbooks and the literature. The genes identified by this search that were not present in ‘Group A’ are referred to as the ‘Group B’ molecules.

In order to ensure that none of these ‘Group B’ molecules were overlooked in the primary search, a secondary PubMed search was undertaken. In the secondary search the ‘Group B’ molecules were individually searched in combination with “ subarachnoid h(a)emorrhage”. Genes of this molecules were searched using official symbol provided by HGNC^28^ and any common alternatives. ‘Group B’ molecules identified in the literature review to have evidence of a role in SAH outcome are referred to as ‘Group C’ molecules. ‘Group D’ molecules represent a more comprehensive set of molecules of interest in SAH outcome (‘Group A’ + ‘Group C’). See Figure 2 for a summary of methodology and molecule groups A-D.

To ensure no molecules were missed, Pubmed IDs from both primary and secondary searches were text-mined using the full-text literature mining gene retrieval and functional analysis tool SciMiner^25^ (http://hurlab.med.und.edu/SciMiner/).

Genes which had not been identified manually were traced back to the original publication to identify whether they were associated with outcome after human or experimental SAH.

## Results

As of 3^rd^ August 2019, 1554 publications were retrieved by the primary Pubmed search. Automated text mining of these publications on SciMiner yielded 295 genes. Manual curation of the publications linked to these genes yielded 43 Group A molecules. A further 15 Group C molecules were identified during the secondary Pubmed search (Figure 2).

The final group of molecules (n=58) relevant to outcome after SAH (Group D = A + C) are shown in Table 1. Table 3 provides further detail on the function of each Group D molecule.

**Table 1.**
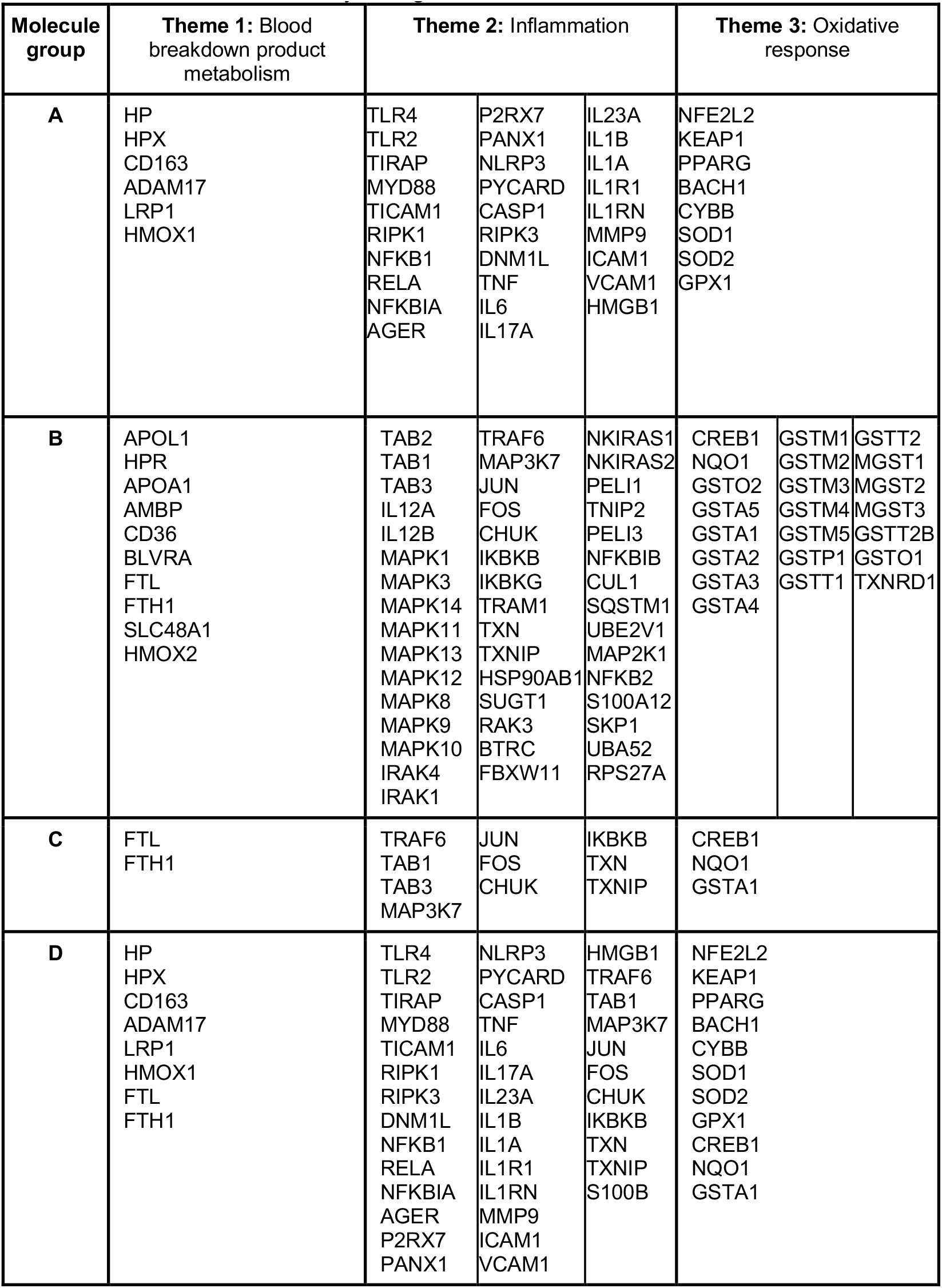
Molecules are denoted by their gene name

**Table 2.**
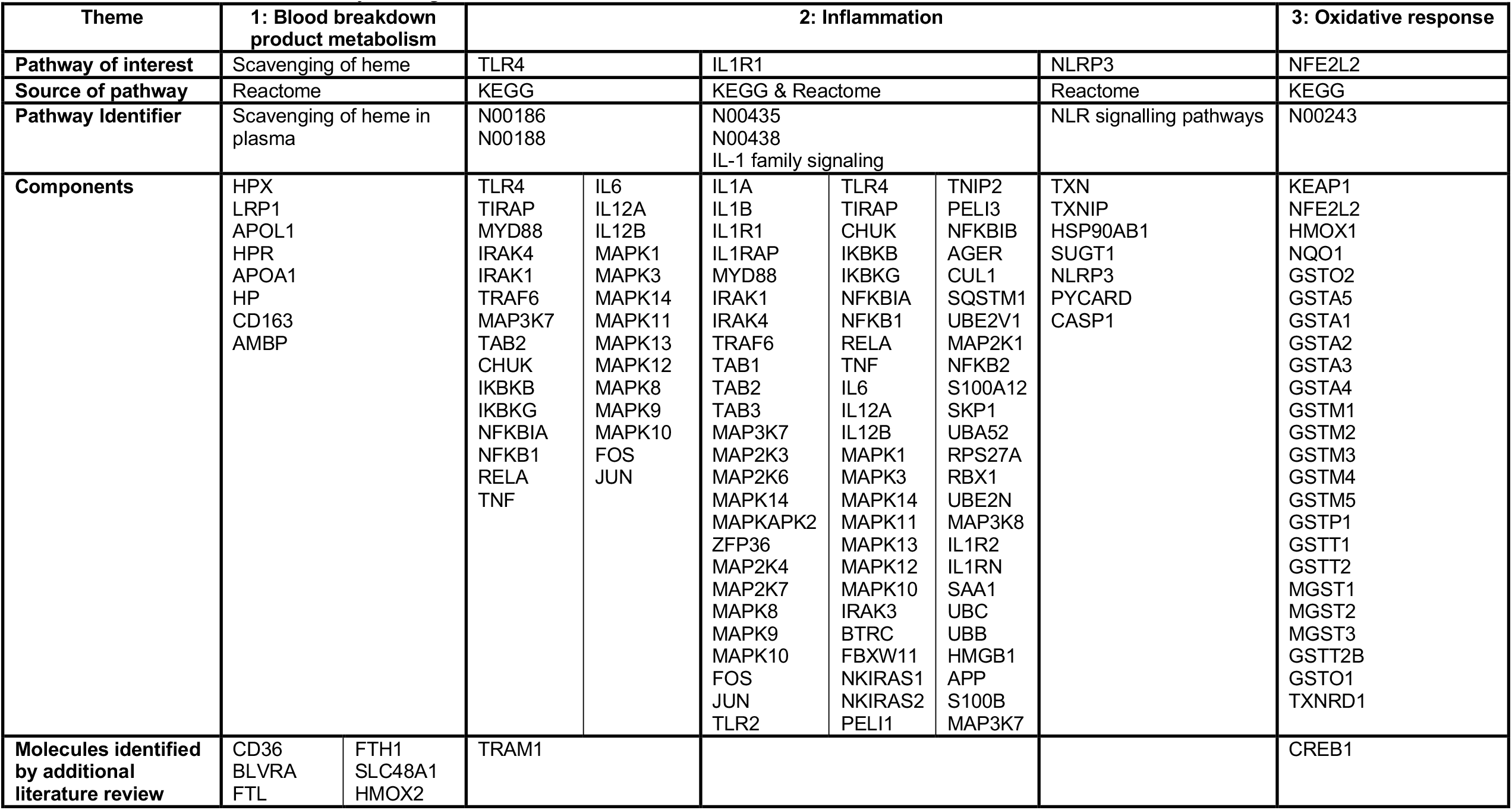
Molecules are denoted by their gene name

**Table 3.**
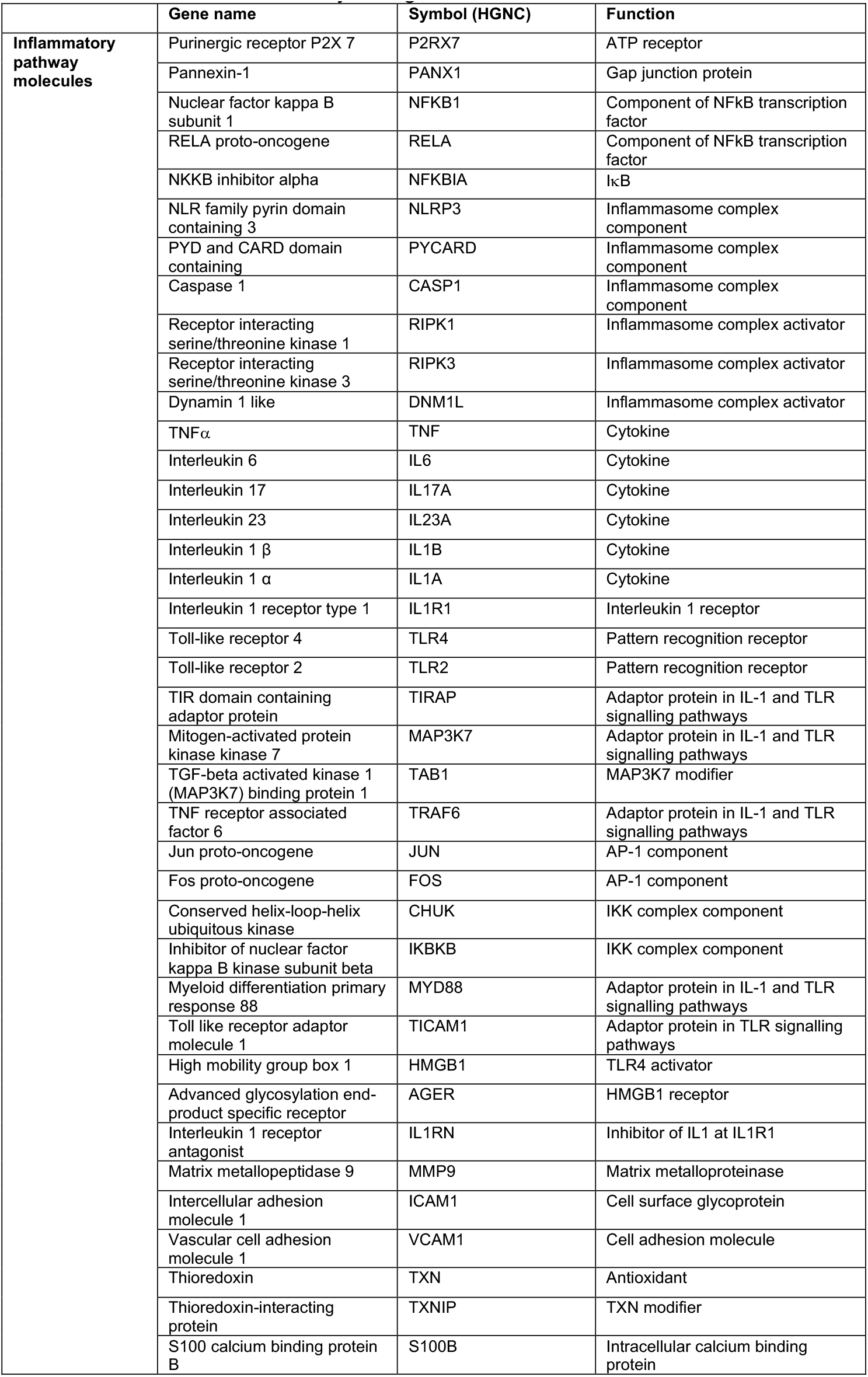
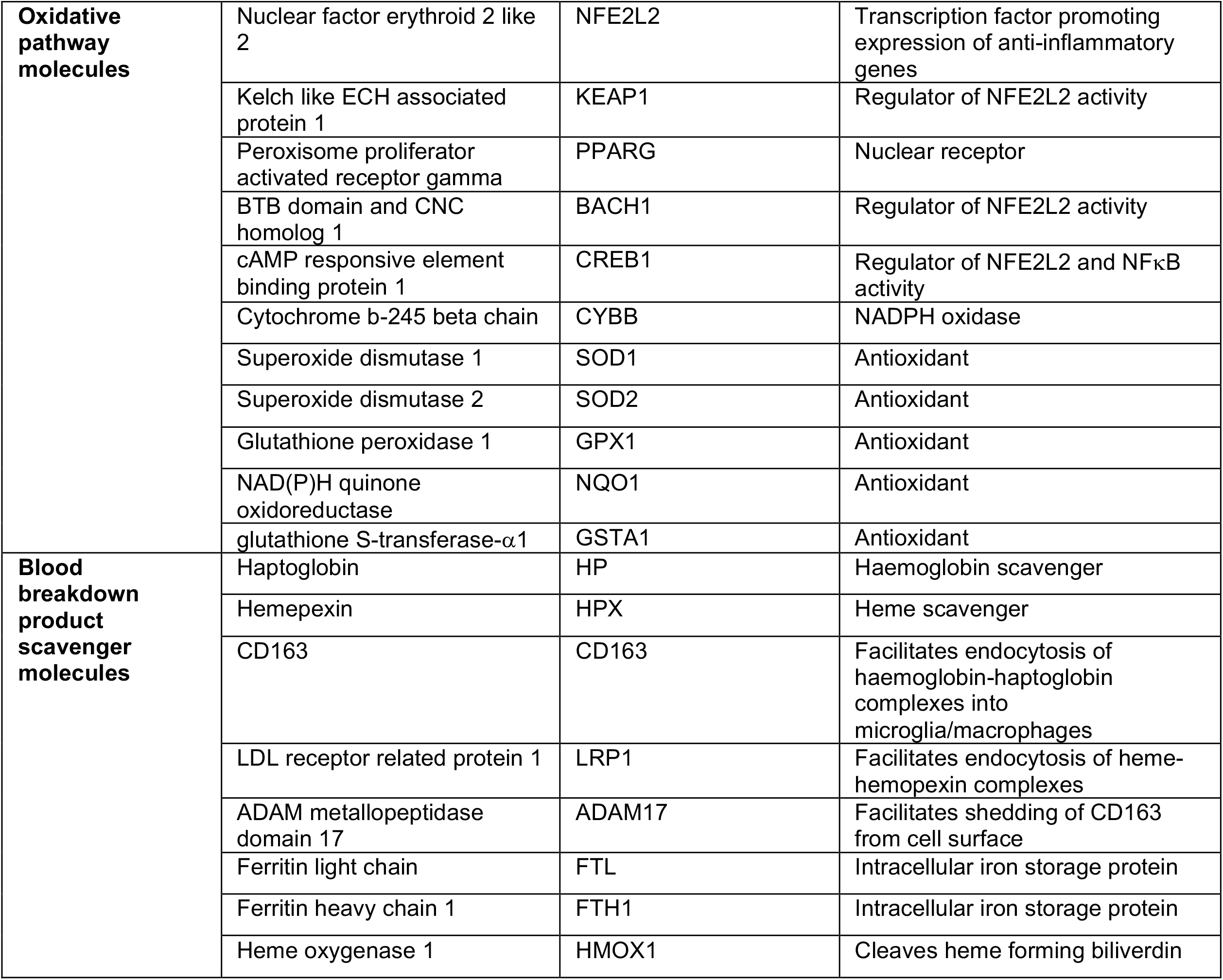
Molecules are denoted by their gene name

## Conclusion

Blood breakdown product metabolism along with inflammatory and oxidative pathways play an important role after SAH. Experimental manipulation and/or genetic variation of molecules in these pathways have been linked to outcome after SAH and they therefore represent a very attractive target for future research studies. Further investigation of these molecules may help inform the mechanisms underlying poor outcome and help develop treatments to mitigate the devastating complications of SAH. It is important to note that these pathways are not independent but intimately linked. Both inflammation and oxidative stress can activate the other pathway, and transcription factors such as Nrf2, PPARγ and NFκB influence both pathways^23,29^. Blood breakdown products can stimulate both inflammation and oxidative stress and their scavengers play an important role to mitigate toxicity by clearing the breakdown products^18,22,30-32^.

In summary, the panel of molecules presented in Table 3 have been identified using a systematic comprehensive search of the literature and will be useful to investigators planning to perform candidate or targeted molecular studies of outcome after SAH. This search will be updated at future intervals.

## Data Availability

This is a systematic review.

